# Predicting the second wave of COVID-19 in Washtenaw County, MI

**DOI:** 10.1101/2020.07.06.20147223

**Authors:** Marissa Renardy, Denise Kirschner

## Abstract

The COVID-19 pandemic has highlighted the patchwork nature of disease epidemics, with infection spread dynamics varying wildly across countries and across states within the US. These heterogeneous patterns are also observed within individual states, with patches of concentrated outbreaks. Data is being generated daily at all of these spatial scales, and answers to questions regarded reopening strategies are desperately needed. Mathematical modeling is useful in exactly these cases, and using modeling at a county scale may be valuable to further predict disease dynamics for the purposes of public health interventions. To explore this issue, we study and predict the spread of COVID-19 in Washtenaw County, MI, the home to University of Michigan, Eastern Michigan University, and Google, as well as serving as a sister city to Detroit, MI where there has been a serious outbreak. Here, we apply a discrete and stochastic network-based modeling framework allowing us to track every individual in the county. In this framework, we construct contact networks based on synthetic population datasets specific for Washtenaw County that are derived from US Census datasets. We assign individuals to households, workplaces, schools, and group quarters (such as prisons). In addition, we assign casual contacts to each individual at random. Using this framework, we explicitly simulate Michigan-specific government-mandated workplace and school closures as well as social distancing measures. We also perform sensitivity analyses to identify key model parameters and mechanisms contributing to the observed disease burden in the three months following the first observed cases on COVID-19 in Michigan. We then consider several scenarios for relaxing restrictions and reopening workplaces to predict what actions would be most prudent. In particular, we consider the effects of 1) different timings for reopening, and 2) different levels of workplace vs. casual contact re-engagement. Through simulations and sensitivity analyses, we explore mechanisms driving magnitude and timing of a second wave of infections upon re-opening. This model can be adapted to other US counties using synthetic population databases and data specific to those regions.

## 1. Introduction

Coronavirus disease 2019 (COVID-19) has spread rapidly throughout the world and the United States (US) since it was declared a global pandemic by the World Health Organization in March 2020 [1]. COVID-19 is an infectious disease caused by the virus SARS-CoV-2, which arose in Wuhan, China, in December 2019 [2]. Studies have shown that COVID-19 is primarily spread from person to person through droplets or direct contact [3, 4]. Common symptoms include fever, cough, and fatigue. Many COVID-19 patients require ventilation and/or intensive care, and the mortality rate among individuals who have tested positive has been estimated to be approximately 7% [5].

Many prevention measures such as a hand washing, social distancing, and quarantine have been instituted, and these have lowered spread significantly (e.g. [6, 7]). The wearing of masks and other personal protective equipment (PPE) has been a controversial issue in the US throughout the pandemic, but data shows that it can stop the spread and lower transmission rates [8]. It is not known how effective these measures will be at continuing to keep case rates low during reopening, and if these measures can prevent a second wave. Recent modeling studies have suggested that relaxing restrictions could have disastrous consequences [9]. Casual contacts between individuals, including going to bars, restaurants, and shops have been associated with driving numbers significantly higher upon reopening in some states like FL, AZ and CA [10].

Mathematical and computational modeling efforts have had an enormous impact on public health policy for the prevention and control of COVID-19 in the US and abroad [11, 12]. For example, the Institute for Health Metrics and Evaluation (IHME) model [13] has been widely cited in the media. A number of model-based forecasts received by the CDC have been made available online [14]. The majority of these models are developed to capture trends and make predictions at the state or nation scale. A variety of modeling methods have been used, including statistical models, ordinary differential equations, partial differential equations, and agent-based models.

While nation-wide and state-wide trends are clearly important, predicting local trends of the COVID-19 pandemic is also of imminent importance given the high heterogeneity (‘patchwork’) of the US and the world. Michigan is one of the hardest hit states in this pandemic in the US so far, with over 58,000 confirmed cases and 5,600 deaths as of June 1, 2020 [15]. In addition to hosting University of Michigan (UM), Washtenaw County is one of the hardest hit Michigan counties outside of the Detroit metropolitan area. In addition, many patients from the city of Detroit have been transferred to UM during the course of the pandemic, raising case numbers in the hospital system. We have thus chosen Washtenaw County, MI as the focus of our study and as a template that can be directly translated to other counties in the US.

In this work, we study COVID-19 in Washtenaw County, MI using a network-based computational model paired with real-world data and synthetic population datasets. The model tracks each individual within the county population in a discrete and stochastic way. We have recently created this model framework and used it to study tuberculosis endemic dynamics within Washtenaw County, MI [16]. Importantly, this model is built on synthetic population datasets built by RTI International that are consistent with US Census datasets [17]. Such synthetic population datasets have been incorporated into other modeling frameworks such as FRED [18], and have been used to study epidemiology of flu-like illnesses (e.g., [19, 20, 21, 22, 23, 24]). Network-based modeling frameworks utilizing these datasets can allow us to simulate realistic scenarios of social interventions since household, school, workplace, and casual contacts are explicitly accounted for every person. This is especially helpful when examining strategies related to workforce re-entry and social distancing.

We are particularly focused on first matching to the model to current Washtenaw COVID-19 datasets, and second, making predictions that can guide re-opening in such a way as to minimize a second wave of infections. We use both uncertainty and sensitivity analyses to consider the effects of 1) different timings for reopening and 2) different levels of workplace vs. casual contact re-engagement. Among other suggestions, we predict that casual contacts between individuals drives the magnitude and timing of a second wave of infections upon re-opening. And thus, we suggest that an abundance of caution should be taken when re-opening social and other non-work-related settings.

## 2. Methods

To study epidemic dynamics of COVID-19, we have taken a discrete stochastic approach, as we believe it provides the most detailed information about the population for the needs of addressing questions about behavior modifications. We outline the model framework, key assumptions, and parameters and how we derived estimates through model calibration and from available datasets through the UM COVID modeling group [25].

### 2.1. Computational model

We have previously developed a network-based model based on synthetic datasets, and used that model to study the dynamics of tuberculosis epidemiology in Washtenaw County, MI as a test case [16]. Briefly, each node in the population network represents an individual in the population under study, and disease transmission events occur in a stochastic fashion through person-to-person contacts occurring within shared households, workplaces, schools, and group quarters, as well as through casual contacts. This modeling framework allows for direct simulation of school and workplace closures and social distancing efforts, as it allows us to individually manipulate transmission dynamics in these different settings.

For our purposes here, we utilize the synthetic population datasets based on US Census data for Washtenaw County as we have done previously [16]; however, we have translated our network model framework from studying tuberculosis to study COVID-19 dynamics. We did this by incorporating the disease progression dynamics used for COVID-19 as developed by Eisenberg et al. [25]. Additionally, we identified essential workplaces in the county so that those businesses would remain open during and after the stay-at-home restrictions. The disease progression framework is shown in Figure 1.

**Figure 1:**
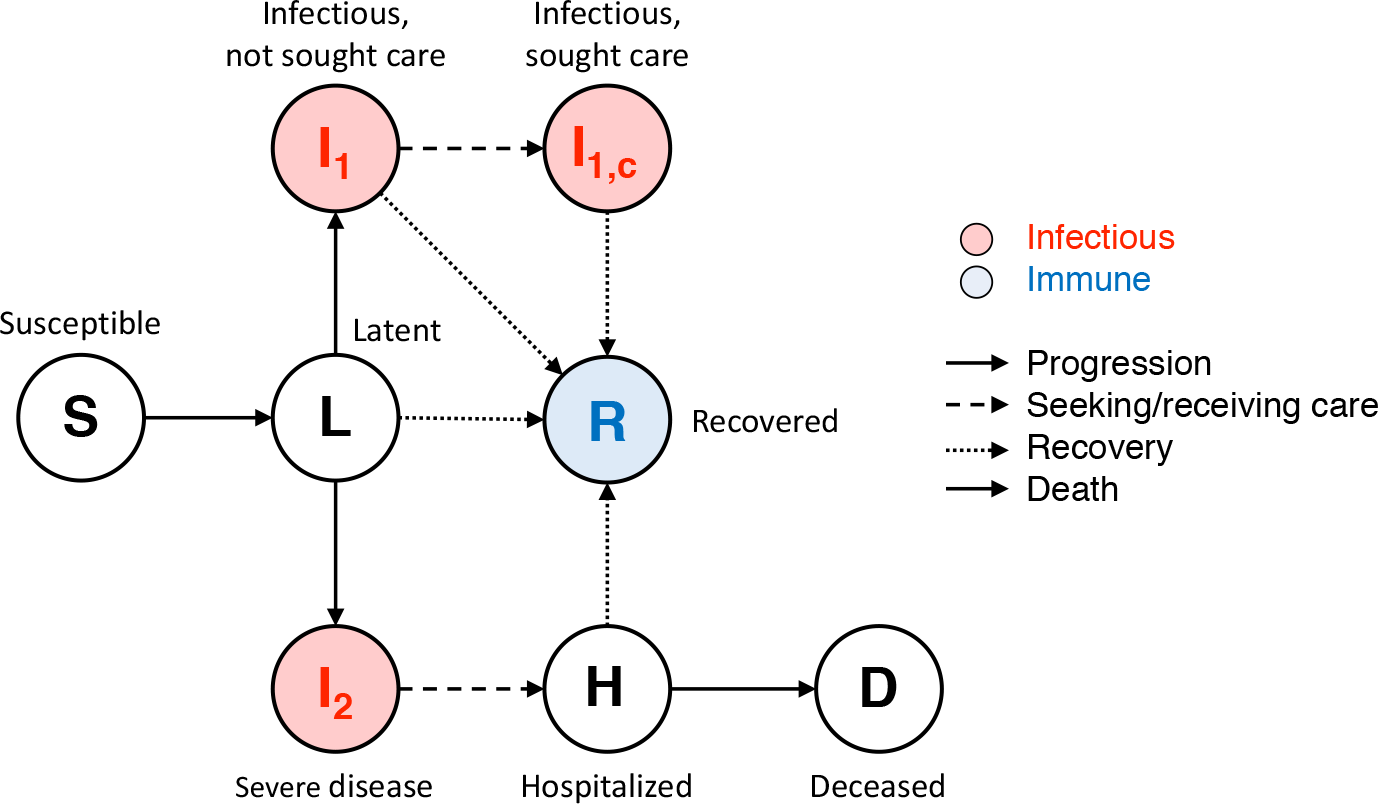
Model flow diagram. Upon exposure, individuals progress from susceptible (S) to latent (L). Latent individuals can develop mild disease (I1), severe disease (I2), or recover (R) without developing disease. Severely infected individuals will become hospitalized (H) and may then die (D) or recover (R). Mildly infected individuals may seek medical care (I1c) before recovering or may recover without seeking care. Recovered individuals (R) are no longer susceptible to infection.

In the network model, susceptible individuals (S) are exposed to coronavirus through contact with an infectious individual (I), with the probability of exposure dependent on the type of contact. For example, we assume that household contacts are more likely to lead to exposure than casual contacts, due to both greater frequencies and duration of contacts. Each type of contact (household, workplace, school, group quarter, and casual) is assigned its own *contact weight*, reflecting the different probabilities of transmission in these different settings. We discuss in Section 2.2 how contact weights are determined. Since the time frame of our simulations is relatively short (less than one year), we use a static network; i.e., we do not include birth, death, or movement between households, workplaces, etc.

After exposure, individuals become latent (L) and then can either recover (R) or progress to becoming infectious (I). Importantly, we assume that latent individuals do not transmit disease. This assumption could be modified in the future as data becomes available on asymptomatic transmission; at the time of this study, however, little is known. Infectious cases are categorized as severe (I2) or non-severe (I1), with ‘severe’ designating cases that will lead to hospitalization. Non-severe cases may either recover without seeking care, or may seek medical care (I1c) before recovery. Severe cases will lead to hospitalization (H), after which individuals may either recover or die (D). Hospitalized patients are no longer able to transmit disease to others in the community, as we assume that they have been effectively isolated. Since data is currently unavailable on transmission within hospital settings, we do not include patient-to-worker or worker-to-worker transmission in hospitals. We simulate protective and isolation measures for individuals who are sick by allowing for reduced infectivity of individuals who have sought care (I1c) or who have severe disease (I2). Finally, we assume that there is protective immunity and that recovered individuals (R) are no longer susceptible to disease, as has been suggested in recent studies [26, 27]. Since our simulations span a relatively short time period, we do not necessarily assume that this immunity is long-lasting; we do assume, however, that it lasts for the time period under study, which is up to nine months for our reopening scenarios.

#### 2.1.1. Simulating closures

To simulate the societal changes imposed during the Michigan state-wide “Stay Home, Stay Safe” order, which took effect on March 24, 2020 and was replaced with relaxed guidelines on June 1, 2020 [28, 29], we made a number of assumptions in the model. For parameter value estimations, see Section 2.2 for details on how we calculated the proportion of workplaces that are deemed essential and the casual contact weight during the stay-at-home order. Additionally, college dorms are removed entirely from the model based on the decisions of UM and other Washtenaw county colleges to suspend in-person classes. Individuals in other types of group quarters, such as prisons and nursing homes, are able to transmit disease within the corresponding group quarter, but are not assigned any casual contacts in the community at large.

To simulate school closures, the school contact weight drops immediately to zero on March 16, 2020. We use a step function for this transition since the closure of public schools occurred on the same day for all elementary and secondary schools across the state of Michigan [30]. To simulate closure of workplaces, the workplace contact weight for non-essential workplaces drops linearly to zero over a period of one week prior to March 24, 2020, when the stay-at-home order went into effect. This one-week ramp-down period reflects increasing closures and precautions implemented during the time between the closure of public schools and the full stay-at-home order, such as restrictions on the use of places of public accommodation [31].

A proportion of workplaces selected at random are deemed “essential”. These were chosen to represent businesses such as grocery stores, pharmacies, hospitals, and others that were not restricted by the stay-at-home order. We reduced the corresponding contact weights associated with essential workplaces to 50% of their baseline values during the stay-at-home order due to mandated precautions such as use of PPE (e.g., masks) and physical distancing. This reduction was chosen arbitrarily and could be modified if data were available on transmission between workers in essential workplaces before and during the stay-at-home order.

To simulate social distancing effects, the casual contact weight decreases linearly to a reduced value that is a fraction of the original over the same one-week period leading up to the stay-at-home order. This fraction is a parameter that is varied and calibrated to match case count data (see Section 2.2).

#### 2.1.2. Other model assumptions

As with any modeling effort, we make assumptions to build and calibrate the model. In addition to assuming the above disease progression framework, we make a number of assumptions about model parameters and the underlying contact network, which we detail here.

Each individual is assigned between 10 and 50 casual contacts, which are randomly selected from the population. Further, individuals in large workplaces, schools, or group quarters (i.e., those with more than 50 members) are assigned between 10 and 50 contacts chosen randomly among the members. In smaller workplaces, schools, and group quarters, all members are assumed to have contact with each other. These limits on numbers of contacts are arbitrary and can be varied as needed or as data are available.

Hospitalization and mortality rates are age-dependent in our model, i.e., they are a function of the age of the person in the population. Other parameters are constant across the population, i.e., they do not vary from individual to individual. We also assume that hospitalized individuals do not transmit disease to the community. An important caveat is that death occurs in our model only after hospitalization, and thus we do not account for deaths happening at home, which have likely been significantly under-reported in hard-hit areas such as New York City [32]. This allows us to better compare with data on confirmed COVID-related deaths, since almost all confirmed deaths occurred in the hospital setting in Washtenaw County; this is possibly due to low testing rates, particularly early in the epidemic. Thus, the predicted numbers of deaths in our simulations are likely to be under-estimates. If we were to model other areas with higher testing and reporting rates, this assumption could be relaxed to allow for deaths at home.

When comparing model simulations with case count datasets [33, 34], we assume that only individuals who have sought medical care could possibly be observed. Thus, we estimate the number of observed cases by taking the sum of cases in the compartments I1c (infectious, sought care) and H (hospitalized), multiplied by their respective reporting rates (see Section 2.2 for how reporting rates are estimated). We assume that reporting rates are constant over time.

Many model parameters have been previously estimated, either from observational data in other COVID-19 [35, 36, 37, 38, 39] or from influenza studies [40], or by using ODE and age-structured models in comparison with case count and death datasets for Washtenaw County. We list these parameters, their estimated values, and references for these estimates are given in Table 1. In our simulations, we set these parameters at their estimated values and vary only the parameters that are unique to our network-based model of COVID-19, with the exception of hospitalization and death parameters, as detailed in Section 2.2.

**Table 1:** Fixed parameter estimates. We show here the parameter estimates for the COVID-19 disease progression framework (Figure 1) together with their units and references that we used to estimate their values

| Parameter | Value | Unit | References |
| --- | --- | --- | --- |
| Basic reproduction number | 2 | people | [41, 3] |
| Incubation period | 5 | days | [35] |
| Infectious period | 7 | days | [37] |
| Mortality fraction among infected individuals | age-dependent | – | [36] |
| Time from symptom onset to death | 18.5 | days | [38] |
| Fraction who are asymptomatic | 0.18 | – | [39] |
| Fraction of symptomatic who will seek care | 0.5 | – | [40] |
| Time to seek care (non-hospital) | 2.5 | days | [41] |
| Fraction of symptomatic who will be hospitalized | age-dependent | – | [36] |
| Time from symptom onset to hospitalization | 11 | days | [38] |
| Duration of hospital stay | 11 | days | [38] |
| Initial proportion of population latent | 1e-4 | – | assumption |
| Initial proportion of population infectious | 1e-5 | – | assumption |

### 2.2. Model calibration

Our model is tailored to specifically study Washtenaw County, MI by building a contact network from a Washtenaw County synthetic population dataset developed by RTI International [42]. This synthetic dataset consists of individuals with sociodemographic features (such as age) who are assigned to households, workplaces, schools, and group quarters such that the population is consistent with county-specific US Census datasets [17].

The network model (Figure 1) is calibrated to match observed COVID-19 datasets on total cumulative cases, hospitalizations, and deaths among Washtenaw County residents between the dates of March 8, 2020 and May 19, 2020. Data are aggregated from Washtenaw County Health Department [33], which includes cases, hospitalizations, and deaths, and also from the New York Times COVID-19 data reports [34], which contains numbers of cases and deaths. Data from Washtenaw County Health Department were collected manually from the web starting on April 6, 2020; however, some earlier time points were recovered using the Internet Archive (https://archive.org). Data from the New York Times are available for every date beginning on March 12, 2020. We assume that cases, hospitalizations, and deaths each have their own constant reporting rate and that these reporting rates are less than one (i.e., cases, hospitalizations, and deaths are under-reported). We sample within the parameter space for parameters that are unique to the network model (such as contact weights and fraction of essential workplaces) using reasonably broad ranges. We typically use Latin hypercube sampling methods for this [43], but here we use Sobol sequences which gives more uniform coverage of the large parameter space [44, 45]. Ranges for contact weight parameters were chosen based on model exploration with the COVID-19 model to establish reasonable upper bounds (data not shown). In addition, since initial exploratory sampling revealed a high number of hospitalizations and a low number of deaths when compared with data, we also varied parameters pertaining to hospitalization and death to obtain the best possible fits. We allow for reduced infectivity of infected individuals who have sought care or who have severe disease, to simulate protective measures. Thus, the list of parameters that we vary for model calibration are: all contact weights, the fraction of workplaces designated as essential, fraction of casual contacts during shutdown, relative infectivity of infected individuals who have sought care, relative infectivity of individuals with severe infection, mortality fraction among infected individuals (age-dependent), fraction of infectious individuals who will be hospitalized (age-dependent), and time to hospitalization. We assume that death and hospitalization rates remain proportional to national rates by age group reported by the CDC [36].

As is typical when we study discrete stochastic models, we explore both epistemic and aleatory uncertainty in the parameter set [43]. This allows us to understand how variations in parameters affect the model outputs (epistemic) and how probabilistic events affect model outputs (aleatory). We sampled 500 parameter sets and performed 5 replications for each parameter set, for a total of 2500 simulations. For each simulation, reporting rates for total cases, hospitalizations, and deaths were individually estimated between 0 and 1 to minimize the respective relative error. Relative errors are measured as 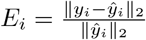 where *y* denotes model output and *ŷ
* denotes observed data, and *i* denotes cases, hospitalizations, or deaths. We define a cost function, as a function of the input parameters **p**, to be the average across replications of the sum of the relative errors.

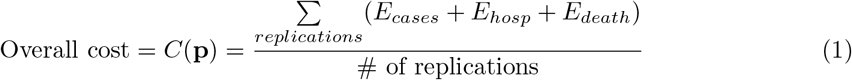

The model was then calibrated by identifying the parameter set **P**_0_ for which cost is best minimized, i.e., **P**_0_ = arg min_**p***∈S*_ *C*(**p**) where *S* is the set of parameter values sampled. Ranges for each of the sampled parameters, as well as the calibrated parameter values **P**_0_, are provided in Table 2.

**Table 2:** Parameter ranges for uncertainty and sensitivity analyses. Minimum and maximum values indicate the ranges used for the initial Sobol sample used to calibrate the model. **P**_0_ denotes the best-fitting parameter values from this sample.

| Parameter | Minimum | Maximum | $\mathbf{P}_0$ |
| --- | --- | --- | --- |
| Household contact weight | 0.8 | 1.2 | 1.16 |
| Group quarter contact weight | 0.8 | 1.2 | 1.13 |
| School contact weight | 0.05 | 0.2 | 0.190 |
| Workplace contact weight | 0.05 | 0.2 | 0.064 |
| Casual contact weight | 0.01 | 0.05 | 0.042 |
| Fraction of essential workplaces | 0.01 | 0.1 | 0.084 |
| Relative infectivity of I1c | 0.5 | 1 | 0.843 |
| Relative infectivity of I2 | 0.5 | 1 | 0.601 |
| Death fraction multiplier | 1 | 2 | 1.77 |
| Hospitalization fraction multiplier | 0.5 | 1 | 0.786 |
| Fraction of casual contacts during shutdown | 0.01 | 0.5 | 0.084 |
| Time to hospitalization (days) | 5.5 | 22 | 6.12 |

Once we identified the parameter set **P**_0_, we defined new parameter ranges to be **P**_0_(1 *±* 0.1). We again performed Sobol sampling to generate a new set of 2500 samples (500 parameter sets with 5 replications each). These simulations yielded model fits that fit well against datasets for Washtenaw County; see Figure 2 for comparison with observed cumulative data. This same parameter range is used to evaluate each reopening scenario.

**Figure 2:**
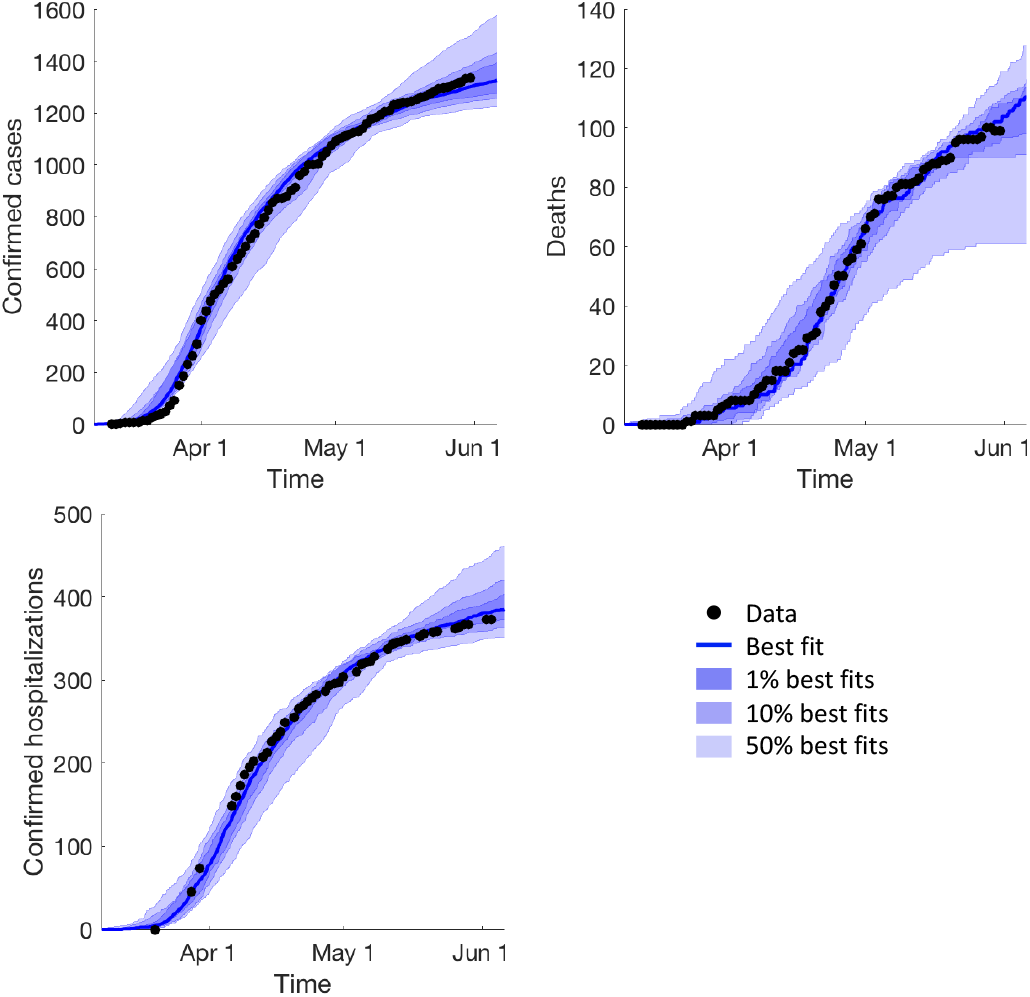
Model fits. Model simulations and observed data are shown for cumulative confirmed cases, deaths, and hospitalizations in Washtenaw County. Black dots indicate observed data, blue lines indicate best fits, and shaded regions indicate the 1%, 10%, and 50% of model runs in the parameter range **P**_0_(1 *±* 0.1) that best fit the data according to the cost function (1).

### 2.3. Uncertainty and sensitivity

We want to determine which system mechanisms, defined via model parameters, can drive different model outputs of interest. To do this, we perform sensitivity analyses for three different model outputs: 1) disease prevalence over time for the first 90 days of simulation under stay-at-home restrictions, 2) the time at which a second peak occurs after reopening, and 3) the peak prevalence after reopening. For outputs 2 and 3 that involve reopening, we use the scenario in which non-essential workplace and casual contact weights return to 50% of normal by July 15, 2020. For output 1, we do not include reopening since the time frame is reasonably short and restrictions were not significantly relaxed within the 90 days following March 8, 2020. For each of these outputs, we compute sensitivities using a set of 2500 simulations as described in Section 2.2.

We quantify parameter sensitivity using partial rank correlation coefficients (PRCC), as this is a nonlinear system and linear correlations may not be appropriate. We follow our usual approach established in [43]. We evaluate significances of PRCC values using a t-test. Since correlations are computed simultaneously for multiple parameters, p-values are corrected using Bonferroni correction. PRCCs and corresponding p-values are computed over time for temporal model outputs, such as numbers of reported cases over time, by calculating them independently at each time step. In our reopening scenarios, we quantify uncertainty in our model predictions by taking the full range of simulations that fall within a 10% error tolerance of the observed data for cases, hospitalizations, and deaths for all dates with at least 20 observations.

### 2.4. Simulating reopening scenarios

We consider two distinct sets of reopening scenarios, one in which we vary the timing of lifting stay-at-home restrictions and one in which we vary the level of casual contact after reopening. In the first set of scenarios, we increase both non-essential workplace and casual contact weights from stay-at-home levels to 50% of normal, occurring over a period of either one, two, or three months beginning on May 15. Here, “normal” refers to the pre-epidemic contact weights defined in Table 2. In the second set of scenarios, we consider the case of reopening over the course of two months from May 15 to July 15. During this time, we increase the non-essential workplace contact weight to 50% of normal while also increasing the saturation level for casual contacts to 50% of normal, 25% of normal, or not increasing casual contacts at all from stay-at-home levels. We do not consider a 100% return to normal since we assume that additional precautions such as physical distancing and using masks or other PPE will still be taken, and will reduce the probability of spreading disease through workplace and casual contacts. These measures have been shown to be effective in reducing transmission [46]. We are assuming here that these measures reduce probability of transmission by 50%; this assumption can be modified in the future as data on effectiveness and compliance becomes available.

Lifting stay-at-home restrictions is simulated by setting contact weights for workplaces and casual contacts equal to functions of the form

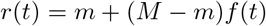

where *m* denotes the contact weight under stay-at-home restrictions, *M* denotes the final contact weight after reopening, and *f* (*t*) is a logistic function that increases from 1% on May 15 to 99% on June 15, July 15, or August 15 depending on the timing of reopening. Figure 3 shows curves of non-essential workplace and casual contact weights over time for each of these scenarios. For each set of reopening scenarios, runs are simulated for a 9-month time frame beginning on March 8, 2020.

**Figure 3:**
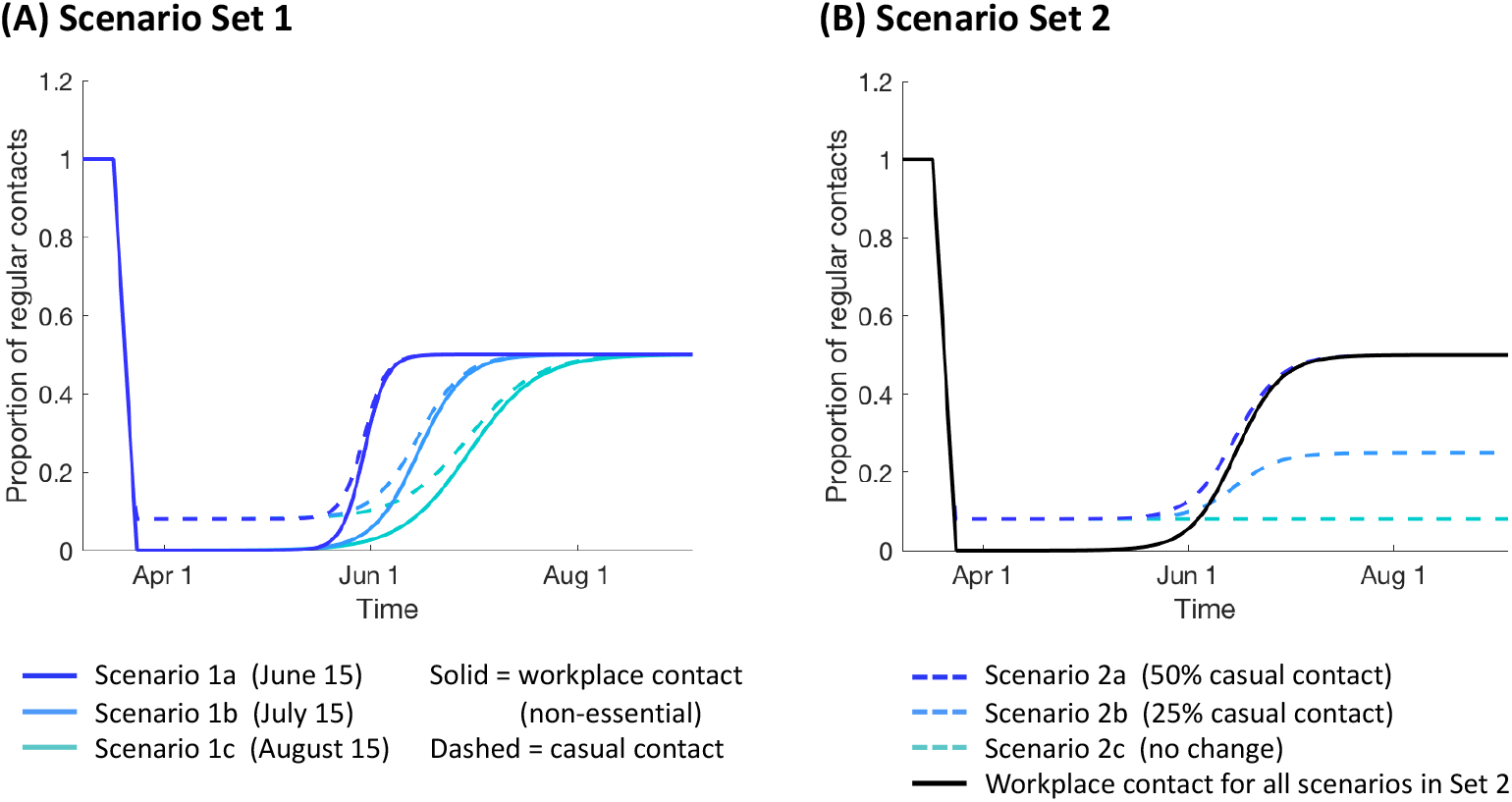
Reopening scenarios. Workplace and casual contact weights over time for two sets of reopening scenarios: 1) varying timing of lifting stay-at-home restrictions (panel A), and 2) varying saturation levels for casual contact (panel B).

## 3. Results

### 3.1. Mechanisms driving disease prevalence

We use sensitivity analysis to evaluate relationships between model inputs (model parameters) and model outputs. Here, we explore the daily disease prevalence for each day of the simulation. This represents the total number of active infections in the population that could possibly be reported at any given time, i.e., the number of cases we would observe with 100% reporting. Due to low levels of COVID-19 testing, although it is improving, true disease prevalence over time is a quantity that cannot currently be empirically measured and can only be inferred through modeling or by making additional assumptions about testing rates.

We perform PRCC analysis using 2500 simulations, consisting of 500 Sobol samples with 5 replications each in the parameter range **P**_0_(1 *±*0.1) as described in Section 2.2. Simulations are run for 90 days beginning on March 8, 2020, which is four days before the first confirmed cases in Washtenaw County. Figure 4 shows the correlation coefficients over time for all parameters that are significant (*p* < 0.01) at any time point during the 90 day window.

**Figure 4:**
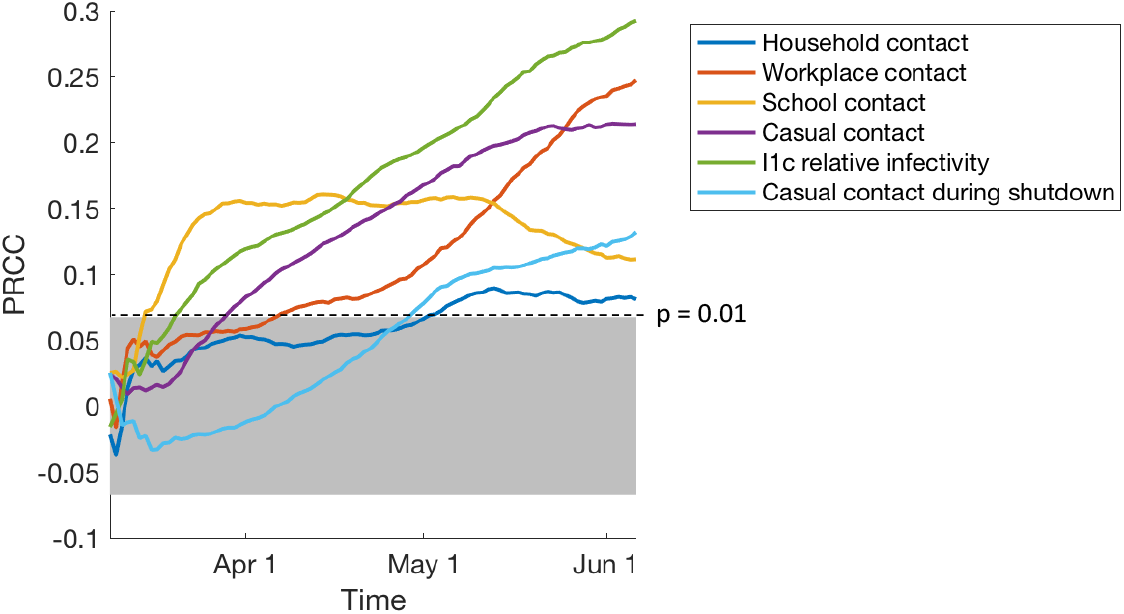
Sensitivity results for disease burden over time predict model mechanisms driving different epidemic outputs. Partial rank correlation coefficient (PRCC) values over time are shown for all parameters that were significant at any time point (*p* < 0.01), using cumulative COVID-19 case count as the model output. Gray shaded area indicates statistical non-significance

The sensitivity analysis predicts that model parameters that are highly correlated (*p* < 0.01) with numbers of daily cases are: contact weights for workplaces, schools, and casual contacts; relative infectivity of individuals who have sought care vs. those who haven’t; and the amount of casual contacts that persist during the stay-at-home order. We find that household contact is less significant than other forms of contact, and only becomes significantly correlated with case counts later in the simulations (after May 1).

These results suggest that uncertainty in the aforementioned parameters leads to significant uncertainty in our model prediction of cumulative numbers of COVID-19 cases. Thus, accurate and reliable estimates for these parameters would enable us to reduce the uncertainty in our model-based predictions for true case load. Further, these parameters represent strong candidates for intervention strategies. Our analysis additionally suggests that reducing contact in workplaces, schools, and casual contacts and encouraging those who are ill to isolate themselves are effective ways of reducing the spread of disease. This aligns with intuition and with the observed flattening of the epidemic curve that has been observed in many regions from precisely these types of interventions [6, 7, 47].

### 3.2. Scenario Set 1: Varied speed of lifting stay-at-home

One of the major questions facing officials regarding reopening is the different speeds for reopening non-essential workplaces and for relaxing social distancing guidelines. While maintaining reduced levels of contact is known to reduce transmission, social and economic costs provide immense pressure to reopen [48]. Thus, it is critical to evaluate the effects of reopening speed on disease burden. To address this question, as discussed in Methods Section 2.4, we consider three scenarios. We increase the contact weights for workplace and casual contacts from stay-at-home levels to 50% of pre-epidemic levels over a period of one, two, or three months starting on May 15, 2020.

Figure 5 shows model projections for each of the three timings considered. We find that decreasing the speed of lifting stay-at-home restrictions only serves to delay the peak of the second wave, but not to decrease its magnitude. Each additional month taken to reach full reopening levels delayed the occurrence of the peak by approximately 18 days on average. In all three scenarios, the median proportion of the population that has been infected (true burden) by early December 2020 is approximately 50%. Therefore, delayed timing affects the timing of the peak, but not its height or the final number of cumulative cases.

**Figure 5:**
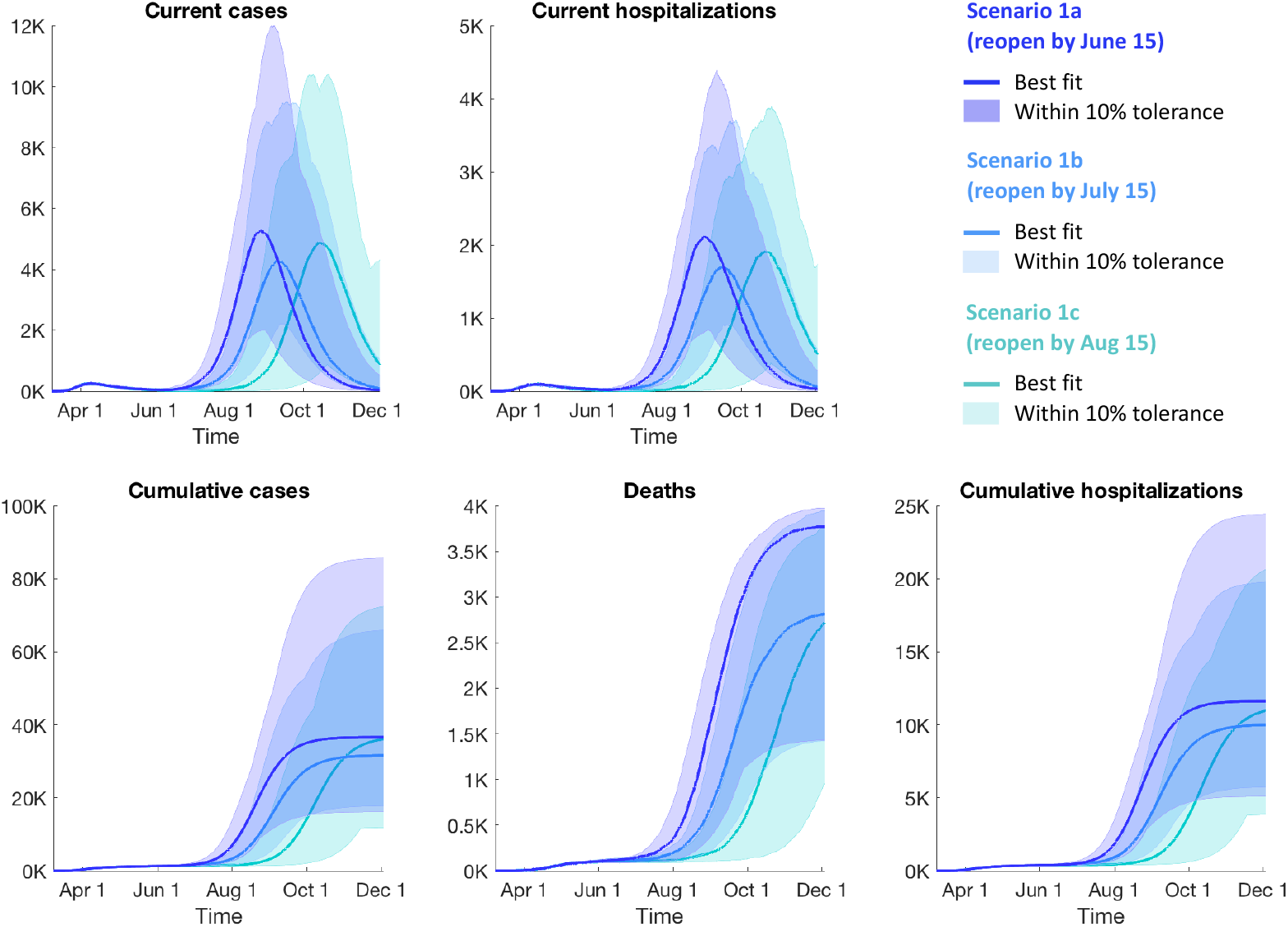
Model projections for Scenario Set 1 (*Varied speed of lifting stay-at-home*) for reported cases, hospitalizations, and deaths. “Current cases” refers to the number of reported infections that are active on a given day. “Cumulative cases” refers to the total number of reported cases that have occurred up until a given date, including recovered cases and deaths. See Section 2.2 for how reporting rates are estimated. Solid lines indicate the simulation that best fit the observed data up to the end of May, and shaded regions indicate the full range of simulations that remained within a 10% error tolerance of all data points with at least 20 observations.

These results indicate that delaying reopening by one or two months is not sufficient to reduce case load at the second peak, but will provide additional time to prepare. The lack of impact on case load appears to be due to a lack of immunity in the population even as reopening occurs over a longer time frame. In particular, by the end of the reopening period (June 15, July 15, and August 15 for scenarios 1a, 1b, and 1c, respectively), the median proportion of the population that is predicted to have become infected is less than 2.5% for each of the three timings, leaving the vast majority of the population still susceptible to infection. Thus, to control case load without an effective vaccine to build individual immunity within the population, we must instead maintain reduced transmission of the virus by maintaining reduced contact.

### 3.3. Scenario Set 2: Varied saturation levels for casual contact

A second question plaguing officials is to what levels to allow a lifting of the Stay-Home, Stay-Safe restrictions. Since slowed reopening has little effect on disease burden during a second wave, as shown above, the degree to which social functions are allowed to reopen and whether PPE and distancing measures should be required will be of utmost importance. To address this question, we consider a second set of scenarios, with a fixed reopening speed, in which casual and workplace contact levels increase from May 15 to July 15. We allow the workplace contact weight to increase to 50% of the normal level, and we vary the final levels of casual contacts to be 50% of normal, 25% of normal, or no change from stay-at-home levels, giving three scenarios in this set. Model predictions for these scenarios are shown in Figure 6.

**Figure 6:**
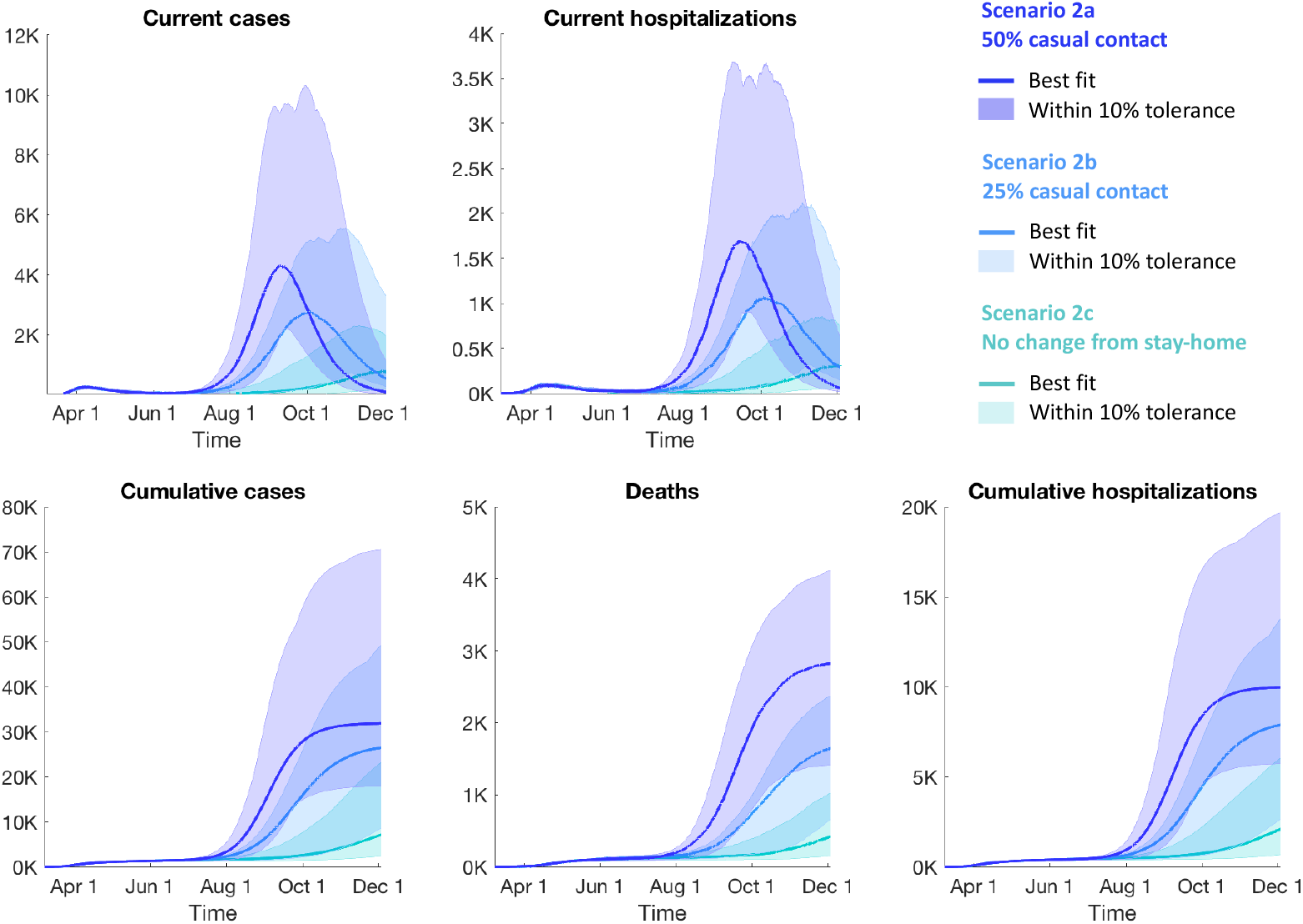
Model projections for Scenario Set 2 (*Varied saturation levels for casual contact*) for reported cases, hospitalizations, and deaths. Solid lines indicate the simulation that best fit the observed data up to the end of May, and shaded regions indicate the full range of simulations that remained within a 10% error tolerance of all data points with at least 20 observations.

The model predicts that decreasing the level of casual contacts (i.e., contacts between people who do not share a household, workplace, school, or group quarter) both delays the second peak and decreases its magnitude by a significant amount. By reducing the final casual contact weight from 50% of normal to 25% of normal, we obtain a 52% reduction in the predicted average peak number of cases and a 34-day delay in the average time to peak. Thus, reducing the amount of casual contacts would both lessen the burden on the local healthcare system (by decreasing the height of the peak) and provide additional time to prepare for the second wave (by delaying the peak). This decrease in contact could be achieved through social distancing and the use of PPE. By further eliminating any increase in casual contact from stay-at-home levels, we obtain an 83% reduction in the predicted average peak number of cases in comparison to the case where casual contacts increase to 50% of normal, and a 64% reduction in comparison to the case where casual contacts increase to 25% of normal. The peak for the case of no increase in casual contact occurs at least 31 days later on average than for the case of casual contacts increasing to 25% of normal; we say “at least” because not all model simulations achieved a peak within the 9 months simulated time frame. Here, averages are computed among all model simulations that remained within a 10% error tolerance of data points with at least 20 observations for cumulative cases, deaths, and hospitalizations.

### 3.4. Mechanisms driving peak timing and prevalence in a second wave

We again utilize sensitivity analysis using PRCC to identify model parameters that are significant for determining both magnitude (based on predicted true burden) and timing of the peak of the second wave of infection that occurs as a result of reopening. We thus use two model outputs for the sensitivity analysis: the number of active cases that have sought or received medical care at the time of the second peak, and the time that the second peak occurs. We performed the analyses presented here for the case of increasing both non-essential workplaces and casual contact weights to 50% of normal levels by July 15, 2020, i.e., Scenario 1a/2b from the above sets of reopening scenarios. Results of the sensitivity analysis are shown in Figure 7.

**Figure 7:**
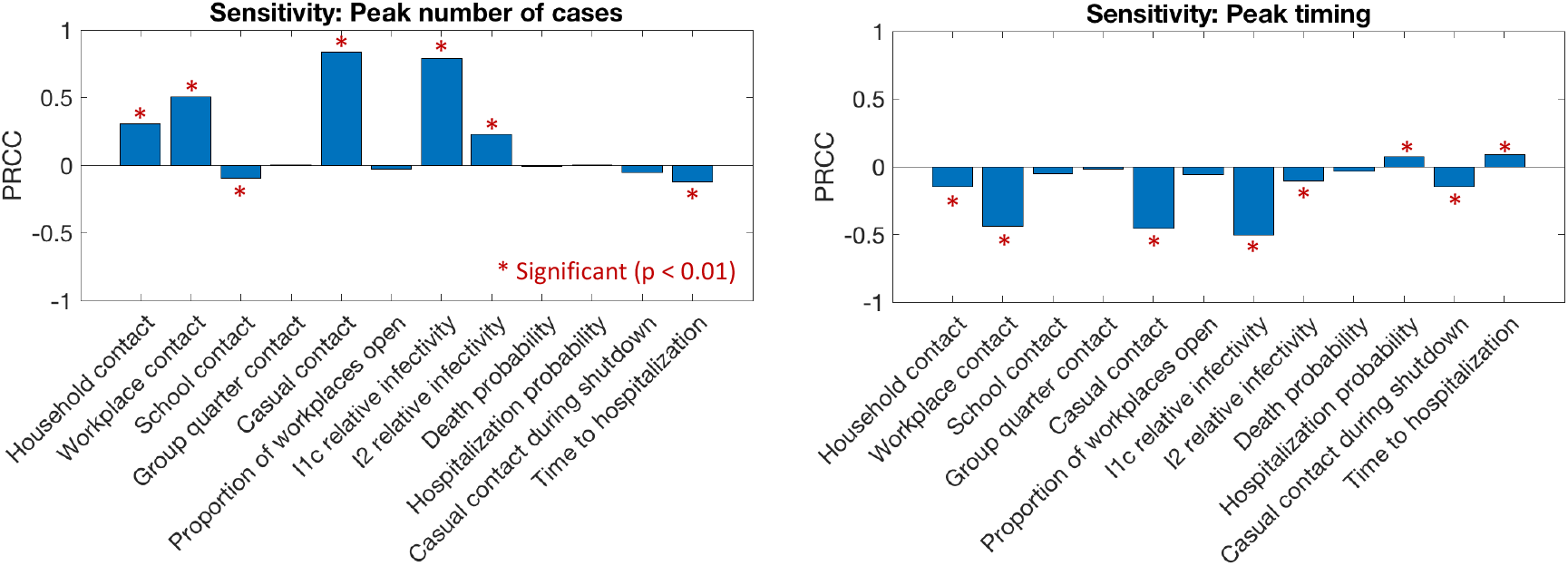
Sensitivity results to identify drivers of case load and timing of the second peak. PRCC results are shown for all varied parameters in the case that workplace and casual contacts increase to 50% by July 15 (Scenario 1b/2a). We consider two model outputs: the number of active cases during the second peak, and the time that the second peak occurs. Asterisks indicate statistical significance (*p* < 0.01).

The most significant parameters for both magnitude and timing of the second peak are work-place contact weight, casual contact weight, and relative infectivity of people who sought care (Figure 7). This suggests that effective strategies for both reducing the peak number of cases and delaying the peak include: (1) reducing workplace and casual contact levels, and (2) encouraging that people who seek care to take protective (such as PPE and distancing) or isolation measures. It further suggests that obtaining accurate data on these parameters would lead to less uncertainty in our model-based predictions for the magnitude and timing of the second peak. We note that there is significant overlap between the most significant parameters for determining the behavior of the second peak and those for determining true case counts during the first wave (see Section 3.1). Other less significant parameters for determining the behavior of the second peak included household contact weights, relative infectivity of people with severe disease, and hospitalization rates.

## 4. Discussion

We predict that delaying reopening on its own is effective only at delaying a second wave, but is ineffective at reducing its magnitude. However, lowering the level of casual contacts after reopening is effective at both delaying the second wave and reducing its magnitude. Further, we predict that casual and workplace contact weights and the reduction in infectivity due to protective measures upon seeking care are the most important factors for determining the magnitude and timing of the peak of a second wave. Thus, social distancing and the use of protective equipment such as masks will be of utmost importance moving forward.

In our model, the contact weights represent relative probabilities of transmission given a certain type of contact. These weights account for frequency and duration of contact as well as other factors that would affect transmission in these settings, such as mask-wearing and social distancing. Thus, the reduced workplace and casual contact weights in our reopening scenarios can be attained either by requiring less frequent or shorter duration of contact or by reducing the probability of transmission through the use of personal protective equipment (PPE) and proper social distancing measures. The contributions of these different approaches to reducing probability of transmission cannot be inferred separately in our model based on fitting to data, since the contact weight represents only the cumulative effect of these efforts. However, if reliable data were available to inform the effects of these interventions on contact weights, effects at the population level for each intervention could be predicted.

In this study, the majority of our model parameters are constant across the population, with the exception of probabilities of hospitalization and mortality, which vary with age. We have not considered here the effects of other sociodemographic features such as race or household income. Since the synthetic population dataset used in our model includes such sociodemographic data, these features could be used in the future to implement subpopulation-specific parameter values. For example, COVID-19 mortality is exceptionally high among African Americans and other communities of color [49, 50]. Model parameters could also be altered to reflect therapeutic interventions and vaccines as they are developed. These interventions could be applied to and affect the population heterogeneously based on age or other features. Age-dependent rates of asymptomatic transmission could also be incorporated into the model when such data becomes available.

Our model can be adapted for use by any other US counties or states, as synthetic population datasets are available for every county and state in the US [42] that can be easily incorporated into the model. Since a realistic contact structure is explicitly built into the model based on US Census data, predictions can be made specific to the social landscape of a specific geographical area. Thus, our model could be used to predict “hotspots” and tailor societal intervention strategies at the county level.

## Data Availability

None

## Acknowledgements

This research was supported by NIH grants R01AI123093 and U01 HL131072 awarded to DEK. The 2010 U.S. Synthetic Population database was created by RTI International, which is funded by the National Institutes of General Medical Sciences (NIGMS). We thank the UM COVID modeling team and Marisa Eisenberg for access and assistance to data and their model. Also to Emily Stoneman, MD in the Division of Infectious Diseases who is the Medical Director of Occupational Health Services an Associate Hospital Epidemiologist in the Department of Infection Prevention and Epidemiology for valued UM datasets used herein.

